# *How Many Patients Do You Need?* Investigating Trial Designs for Anti-Seizure Treatment in Acute Brain Injury Patients

**DOI:** 10.1101/2023.08.21.23294339

**Authors:** Harsh Parikh, Haoqi Sun, Rajesh Amerineni, Eric S. Rosenthal, Alexander Volfovsky, Cynthia Rudin, M. Brandon Westover, Sahar F. Zafar

## Abstract

**Objectives:** Epileptiform activity (EA) worsens outcomes in patients with acute brain injuries (e.g., aneurysmal subarachnoid hemorrhage [aSAH]). Randomized trials (RCTs) assessing anti-seizure interventions are needed. Due to scant drug efficacy data and ethical reservations with placebo utilization, RCTs are lacking or hindered by design constraints. We used a pharmacological model-guided simulator to design and determine feasibility of RCTs evaluating EA treatment.

**Methods:** In a single-center cohort of adults (age >18) with aSAH and EA, we employed a mechanistic pharmacokinetic-pharmacodynamic framework to model treatment response using observational data. We subsequently simulated RCTs for levetiracetam and propofol, each with three treatment arms mirroring clinical practice and an additional placebo arm. Using our framework we simulated EA trajectories across treatment arms. We predicted discharge modified Rankin Scale as a function of baseline covariates, EA burden, and drug doses using a double machine learning model learned from observational data. Differences in outcomes across arms were used to estimate the required sample size.

**Results:** Sample sizes ranged from 500 for levetiracetam 7 mg/kg vs placebo, to >4000 for levetiracetam 15 vs. 7 mg/kg to achieve 80% power (5% type I error). For propofol 1mg/kg/hr vs. placebo 1200 participants were needed. Simulations comparing propofol at varying doses did not reach 80% power even at samples >1200.

**Interpretation:** Our simulations using drug efficacy show sample sizes are infeasible, even for potentially unethical placebo-control trials. We highlight the strength of simulations with observational data to inform the null hypotheses and assess feasibility of future trials of EA treatment.

## Introduction

Epileptiform activity (EA), characterized by seizures, periodic and rhythmic patterns, is strongly linked to higher mortality rates and poor functional outcomes in patients with acute brain injuries (including trauma, ischemic and hemorrhagic stroke).^1-4^ Up to 50% of patients with acute brain injury undergoing continuous electroencephalography (EEG) exhibit EA.^5-9^ Although anti-seizure medications (ASMs) such as levetiracetam or propofol are commonly prescribed to manage EA in patients with acute brain injuries, there is limited evidence that such treatment enhances long-term outcomes.^4,8,10-13^

Randomized controlled trials (RCT) are considered the gold-standard approach to assess the effectiveness of clinical treatments and establish evidence-based treatment guidelines.^14^ However, conducting RCTs for EEG-guided anti-epileptiform treatment faces challenges stemming from the variability in drug response among patients. Individuals may need different dosage regimens and treatment durations, while clear definitions of treatment targets and endpoints are ill defined. Moreover, standardizing the timing of EEG initiation and determining the time to randomization can be challenging due to the variable presentation time from ictus. Lastly, randomizing individuals into treatments that are considered likely to be worse than standard of care is problematic due to ethical considerations. To design a robust trial, it is imperative to meticulously consider these factors.

One of the key aspects of a study design is the determination of a reasonable sample size via a power analysis.^15^ Sample size selection in prior RCTs on EEG-guided anti-seizure treatment were not based on realistic power analysis calculations. Specifically, they did not account for the above factors due to the limited availability of data on pharmacodynamic heterogeneity and poor-quality data on outcomes. This resulted in unrealistic null distributions and consequently underestimation of the required sample size.^16,17^ To address these challenges, we propose leveraging observational data in a simulation-aided design of a randomized experiment and power analysis. This simulation-aided approach assists in establishing appropriate sample sizes, optimizing the trial design and improving the reliability of future estimates.^18-20^

In this study, we employed mechanistic pharmacological model-guided simulations to assess the feasibility of a trial investigating the effectiveness of anti-seizure medication (ASM) treatment for EA in a subgroup of acute brain injury patients with aneurysmal subarachnoid hemorrhage (aSAH). The target endpoint for these trials was functional outcomes at discharge as measured by the modified Rankin Scale (mRS). Our analysis focuses on evaluating the impact of trial design and drug efficacy in reducing EA burden on the required sample size needed to achieve sufficient statistical power. To accomplish this, we developed a simulator that incorporated real EEG and ASM data from aSAH patients, enabling us to understand and simulate patients’ epileptiform activity trajectories during their hospitalization, including the interactions between EEG and ASM. Using the trained model, we conducted simulated RCTs, varying the ASM doses, to investigate their effects on the modified Rankin Scale.

## Methods

We developed a framework to leverage observational data to inform the efficient design of experiments in clinically and physiologically complex scenarios (see Figure 1). Specifically, we studied sequential ASM treatment regimes, where administered treatments not only affect the short-term state (i.e. EEG findings) but also the long-term outcome of a patient (e.g., discharge modified Rankin Score [mRS]), while at the same time capturing patient-level heterogeneity in response to treatment. We designed a simulated randomized control trial that used a mechanistic simulator and long-term effect estimates learned using observational data. This design allowed us to perform power analyses and compare various choices of treatment arms and outcomes.

**Figure 1.**
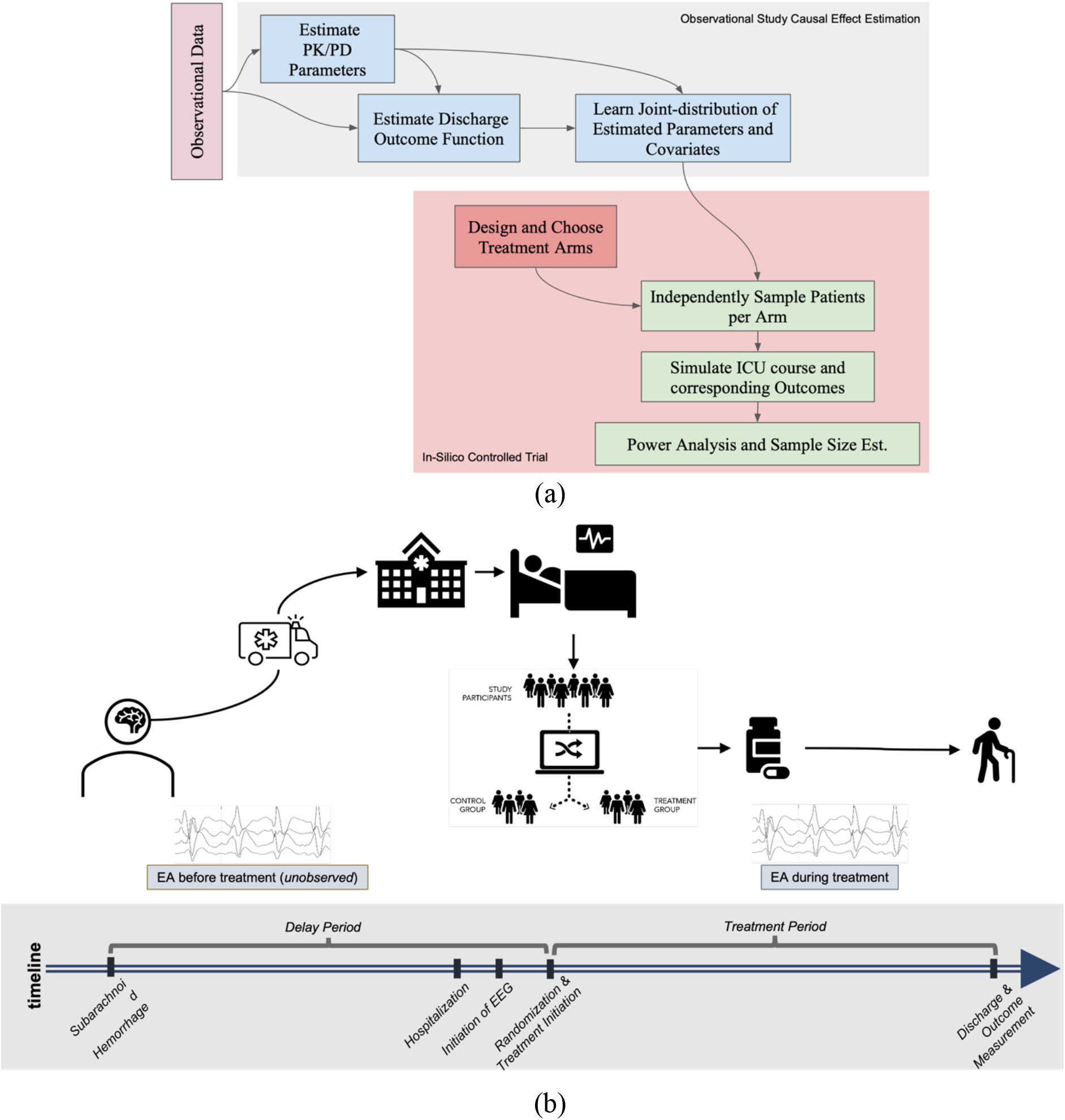
The overall analysis framework and a typical patient’s timeline. (a) The analysis framework consists primarily of two parts -- estimation using observational data (shown in grey) and simulated RCTs (shown in red). (b) The schematic shows the timeline of a typical patient participating in a prototypical RCT.

### Patient Cohort

The observational study was approved by the Institutional Review Boards of the Massachusetts General Hospital and Duke University. Informed consent was not required. The EEG recordings and clinical and demographic variables were extracted from a retrospective database of adult (age ≥ 18 years) aSAH patients admitted to Massachusetts General Hospital between 2012 and 2017. From the initial database of 136 patients with aSAH, we considered a subset of 48 patients who underwent more than 24 hours of EEG and had EA during monitoring. Here, EA was defined as seizures, generalized and lateralized periodic discharges (GPDs, LPDs), and lateralized rhythmic delta activity (LRDA), as all of these are highly associated with seizures and are frequently treated with anti-seizure medications in clinical practice.^1,4,8,21^ Generalized rhythmic delta activity (GRDA) was excluded from our definition of EA as it is a more benign pattern that is not associated with seizures.^8^ Clinical and demographic variables for the selected patients are shown in Table 1. The median time from admission to EEG initiation was 24 hours. Discharge outcome was measured using the modified Rankin Scale. We dichotomized outcomes as good (mRS < 4) and poor (mRS ≥ 4). The data and software to reproduce these findings are available in a publicly accessible repository at bdsp.io.

**Table 1.**
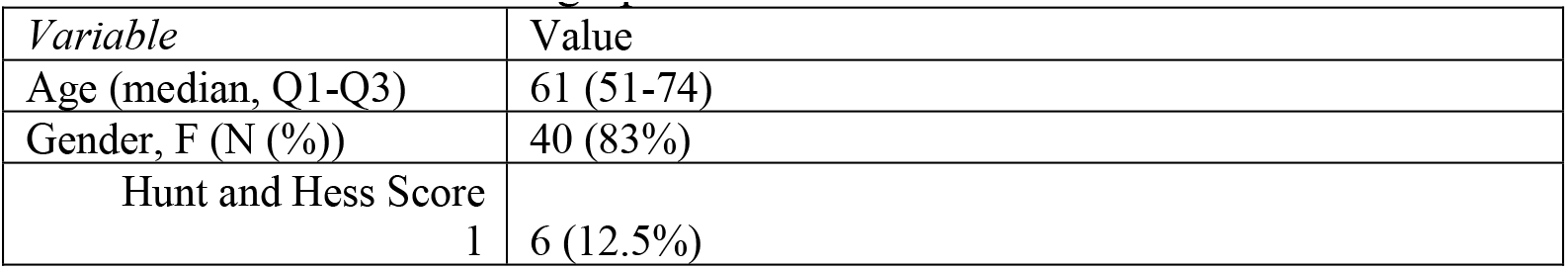
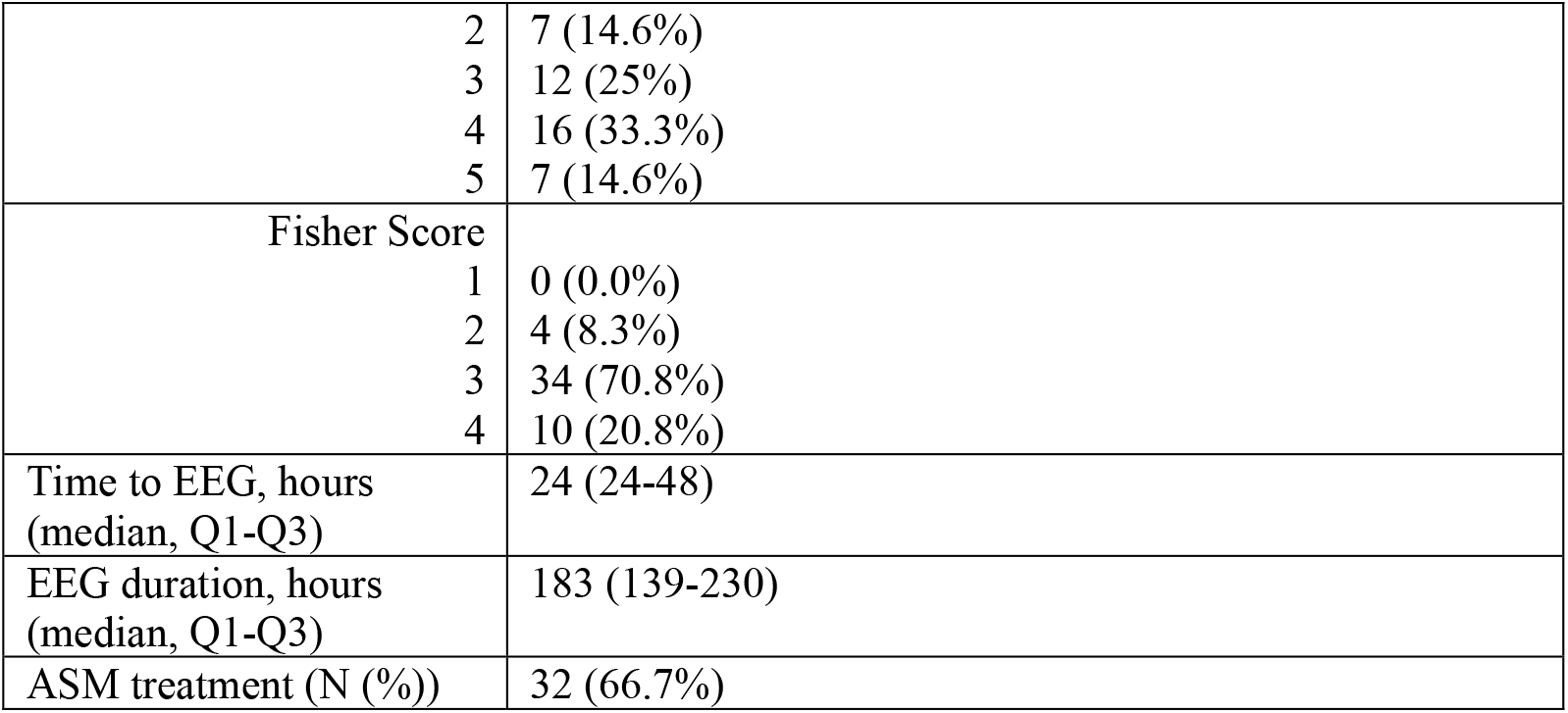
Clinical and Demographic variables.

| Variable | Value |
| --- | --- |
| Age (median, Q1-Q3) | 61 (51-74) |
| Gender, F (N (%)) | 40 (83%) |
| Hunt and Hess Score |  |
| 1 | 6 (12.5%) |

|  |  |  |
| --- | --- | --- |
|  | 2 | 7 (14.6%) |
|  | 3 | 12 (25%) |
|  | 4 | 16 (33.3%) |
|  | 5 | 7 (14.6%) |
| Fisher Score |  |  |
|  | 1 | 0 (0.0%) |
|  | 2 | 4 (8.3%) |
|  | 3 | 34 (70.8%) |
|  | 4 | 10 (20.8%) |
| Time to EEG, hours<br>(median, Q1-Q3) |  | 24 (24-48) |
| EEG duration, hours<br>(median, Q1-Q3) |  | 183 (139-230) |
| ASM treatment (N (%)) |  | 32 (66.7%) |

### EA Burden Estimation

Increasing EA burden is associated with worse outcomes not only in aSAH patients, but across a wide spectrum of acute brain injury patients.^1,2,4^ We therefore examined the interaction of EA burden and ASM treatment with outcomes. To measure EA burden in our cohort of aSAH we followed these steps: First, a previously developed convolutional neural network (CNN) classifier was used to label every consecutive 2-second segment of cEEG data as having one of the EA patterns (seizure, GPD, LPD, LRDA) or not.^22-24^ Then, the EA burden time series was defined as the proportion of 2-second segments having EA in a moving non-overlapping 10 minute window.^1,2^ We used a clinically meaningful summary to quantify EA burden as E_max_: the maximum EA fraction among six-hour sliding windows. This measure of EA burden was selected based on our prior work demonstrating that the maximum EA burden within a sliding 6-hour window has a significant negative effect on discharge neurologic outcome.^2^

### Anti-Seizure Medication and PK/PD Modeling

We modeled the short-term effect of ASMs on EA using a mechanistic pharmacokinetics-pharmacodynamics (PK/PD) model. Mechanistic PK/PD models are based on biological processes and the parameters have physical or biological interpretation. Estimating PK/PD parameters for each patient allowed us account for patient heterogeneity across the cohort, which is important in our application. Thus, using mechanistic models guarantees interpretability of the estimates and is well-suited for our application as these models require fewer data to calibrate compared to empirical (nonparametric) statistical models. We used a one-compartment PK model to estimate the concentration of drugs for each patient in the cohort.^25^ Further, we used Hill’s PD model to estimate the short-term effectiveness of the ASMs in reducing the EA burden.^26^ We estimated the drug concentration necessary to reduce EA burden in a 10-minute window by 50% from the maximum level and the steepness of the effect-onset curve (Hill’s coefficient).^27^ We estimated the PK/PD parameters using the observed EA burden and ASM time series from each patient to account for inter-patient heterogeneity. Our estimation method minimizes the mean-squared error between the simulated and observed EA burden trajectories.

### Discharge Outcome Modeling

Following our result showing E_max_ in a 6-hour sliding window negatively impacts outcomes^2^, we focused on modeling the outcome (mRS) as a function of E_max_ and average ASM concentration during treatment. We adapted the doubly robust machine learning causal effect estimator^21^ using gradient boosting trees to learn discharge outcomes as a function of E_max_ and ASMs.^2^ We adjusted for all baseline features (described in Table 1) and their PK/PD parameters as confounders.

### Simulated Controlled Trials

We designed trials to estimate the effectiveness of varying treatment regimens for a commonly prescribed ASM, levetiracetam, and a commonly prescribed anesthetic, propofol. We used the models learned using the observational data (as described in the previous section) to simulate RCTs by generating random samples of patients’ EA burden over time (as a function of the ASM treatment regime) as well as discharge outcomes (as a function of EA burden and ASM exposure). We analyzed how the required sample size and associated power varied with the choice of treatment arms and outcomes defined below. We evaluated the outcome measure of discharge neurologic status measured using the modified Rankin Scale (mRS) dichotomized as good (mRS 0-3) and poor (mRS 4-6) outcome.^2^

### Treatment arms

For each of the two drugs, we designed three treatment arms based on the drug dose plus standard care and a fourth placebo arm. For levetiracetam, we used the following three doses (administered every 8 hours): (i) 15 mg/kg, (ii) 7mg/kg, (iii) 3mg/kg, and (iv) placebo with standard care. Similarly, for propofol the treatment arms were: (i) 1mg/kg/hr, (ii) 0.5 mg/kg/hr, 0.25 mg/kg/hr, and (iv) placebo with standard care. For both drugs, the second treatment dose (7mg/kg for levetiracetam and 0.6 mg/kg/hr for propofol) were chosen based on the median doses used in the observational data.

We simulated RCTs with varying sample sizes using our cohort of 48 patients. For each sampled patient, we simulated a timeline akin to the real-world scenario (as shown in Figure 1b). The baseline covariates such as demographics and medical history were measured at time of hospitalization. We considered the first EA measurement as t=0. After initial measurement of EA, the patient was randomized into one of the four treatment arms for each drug, in addition to the standard of care as per the observed data Simulated outcomes were measured at hospital discharge.

We compared pairwise outcomes for each treatment arm and computed the power of the analysis. Thus, varying the sample size and re-performing the analysis allowed us to identify the smallest sample needed to achieve a power of more than 80% for each treatment arm and outcome, with effect sizes as observed in our data, using a 2-sided T-test.

## Results

We estimated power curves for 3 combinations of treatment arms for both levetiracetam and propofol (see Figure 3). We found that comparing levetiracetam or propofol to placebo yielded the maximum power for a given sample size, as opposed to comparing different treatment intensities. For levetiracetam, a sample size of at least 500 patients per treatment arm is needed to achieve power of more than 80%, whereas for propofol, at least 600 patients are needed per treatment arm. The need for large sample sizes can partially be attributed to the relatively small effects of these drugs on discharge outcome mRS. For both drugs of interest, Figure 2 shows the expected discharge outcomes (measured as binarized mRS) for each treatment arm estimated using the clinical trial simulation. For levetiracetam, we found that a reduction of the drug dose from 15mg/kg to 7mg/kg improves the outcome – this may be related to the potential negative side effects of a high drug dose. However, further reduction in levetiracetam dose from 7mg/kg to 3mg/kg worsened outcomes, due to the impact of untreated higher EA burden. Interestingly, for propofol, we found no or minimal difference in outcomes between the highest (1mg/kg/hr) vs. lowest dose (0.25mg/kg/hr) treatment arms. This could potentially be explained by the similar impact of propofol on EA at all three doses, and that the highest propofol dose (1mg/kg/hr) used in this cohort was also a low dose leading to minimal side effects.

**Figure 2.**
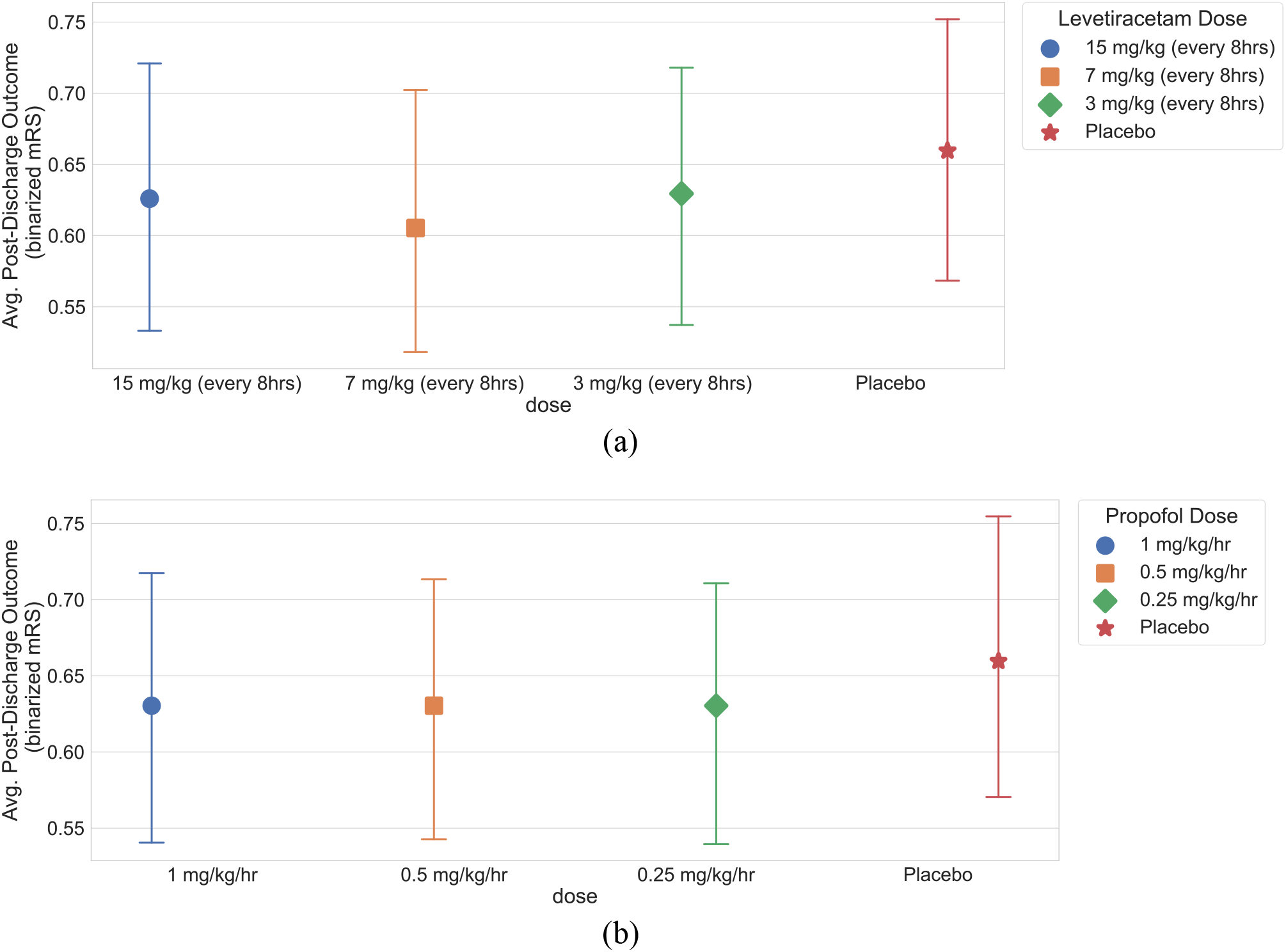
Expected post-discharge outcomes (binarized mRS) under each treatment arm for (a) levetiracetam and (b) propofol estimated using the observational data. The points show mean point estimate and the error bars show standard error.

**Figure 3.**
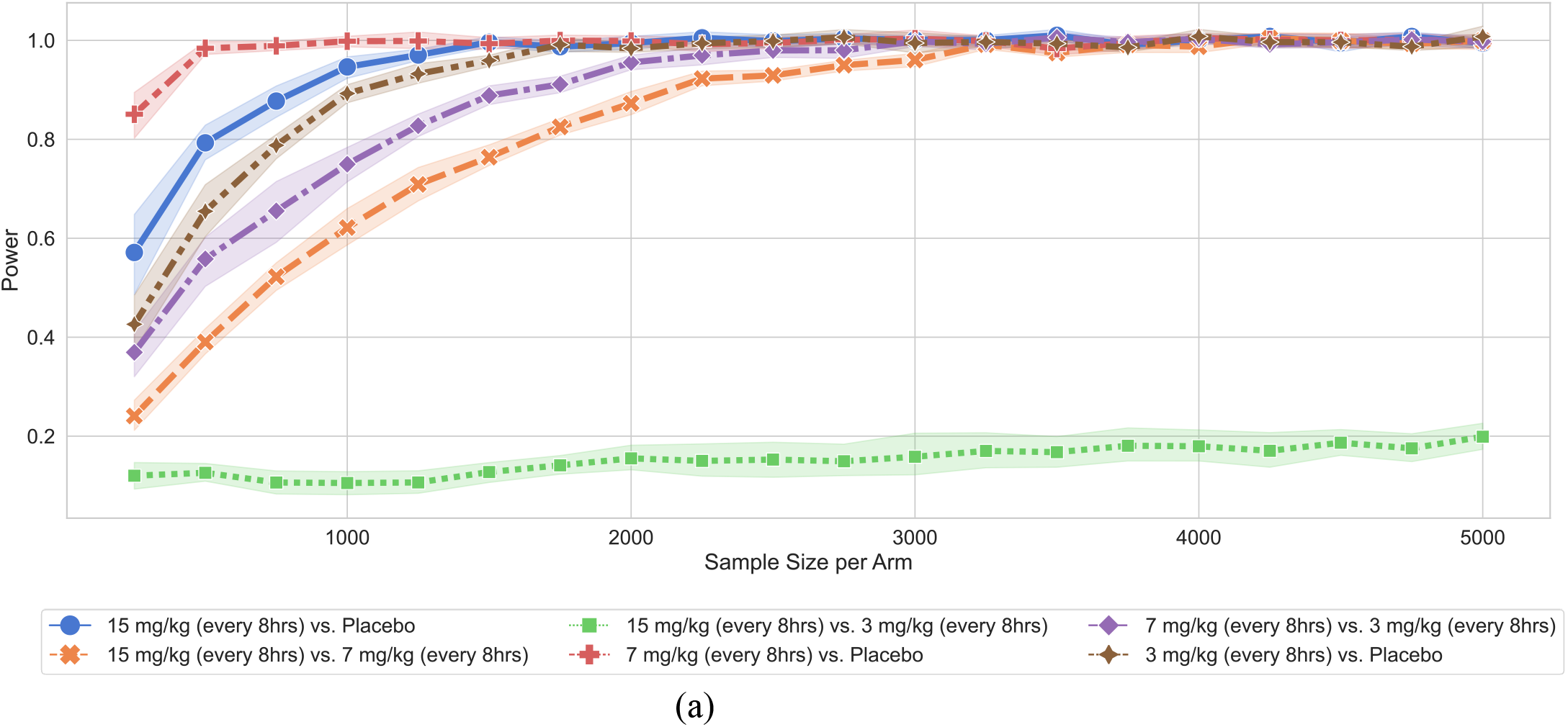
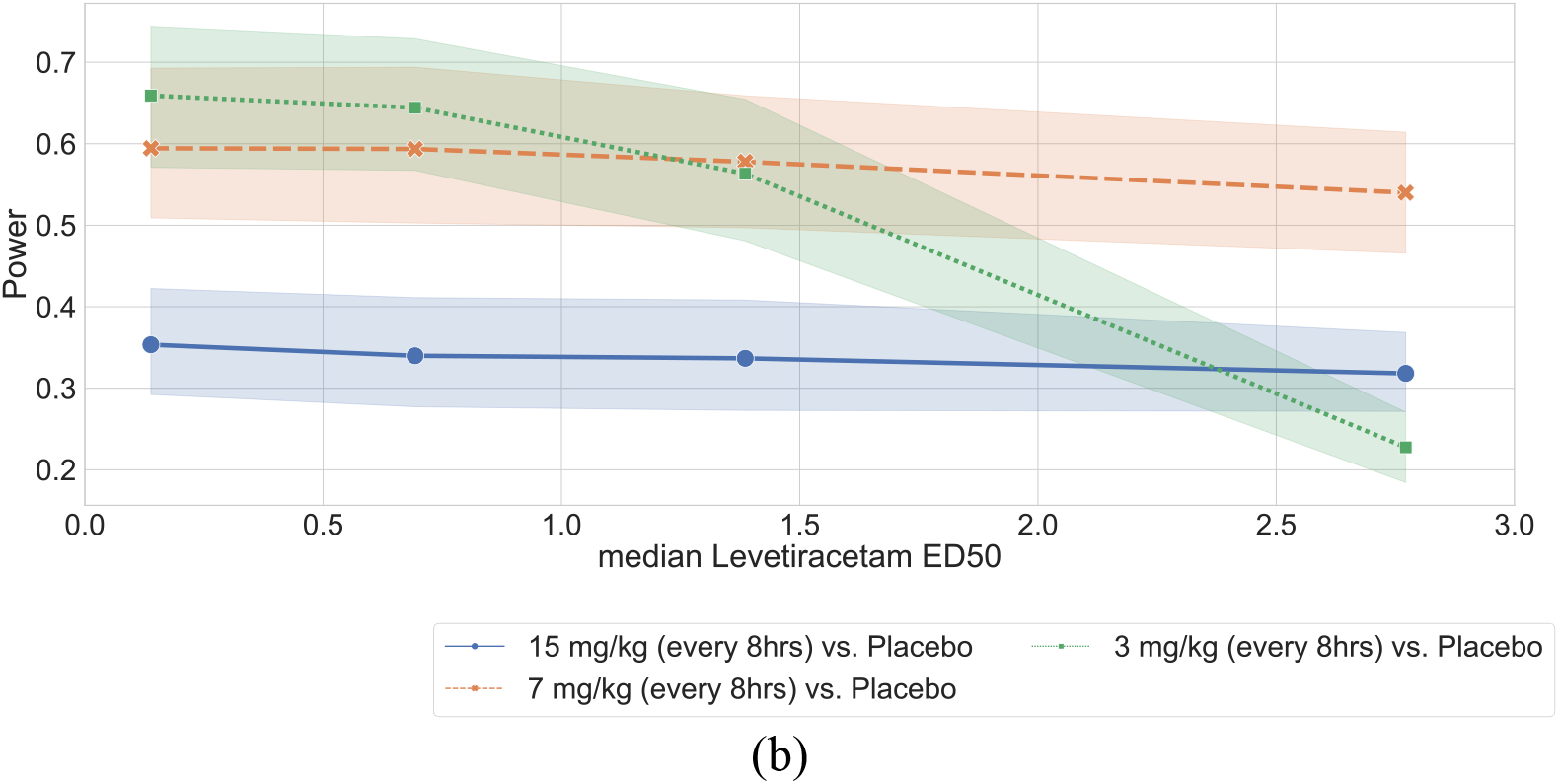
Power analysis and sample calculation for levetiracetam. (a) We estimate the sample size for 80% power calculated for binary combinations of treatment arms on binarized discharge mRS scores. (b) For a sample size of 100 units per treatment arm, we change the ED50 for levetiracetam to identify the drug-effect size at which the treatment effect is detected with 80% power on the discharge outcome.

**Figure 4.**
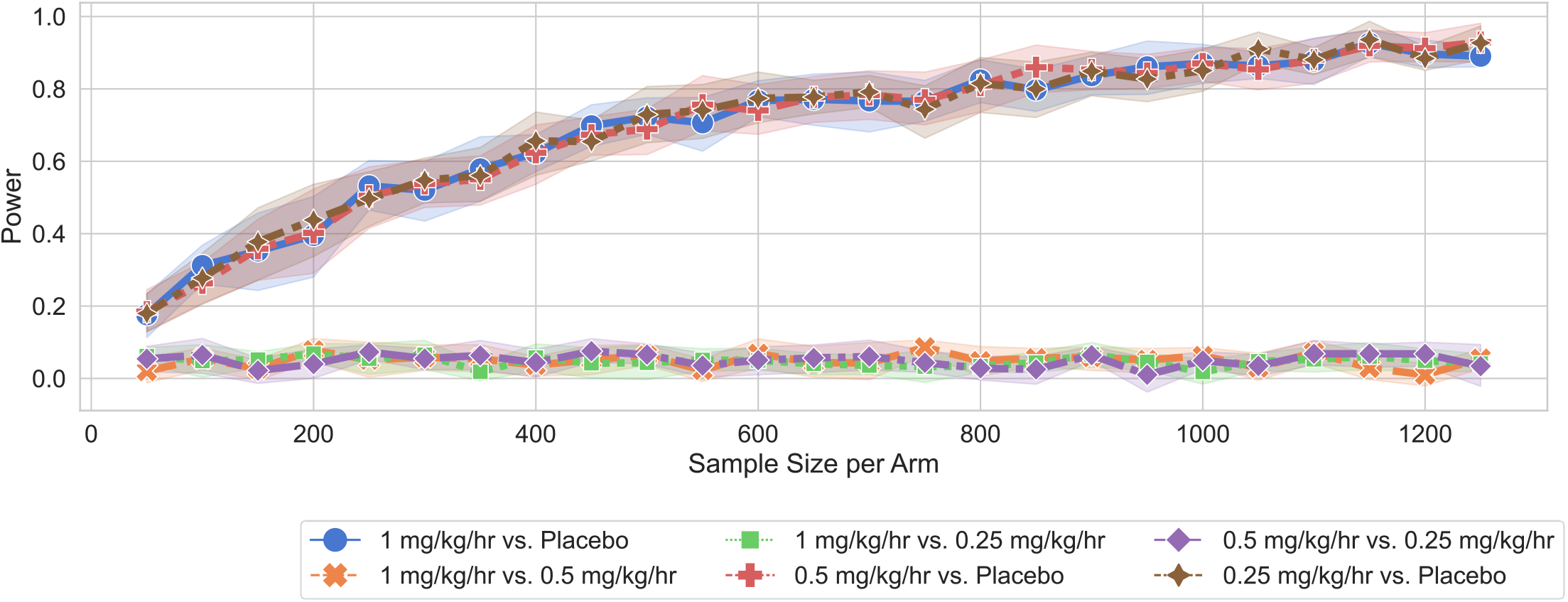
Power analysis and sample calculation for propofol. We estimate the sample size for 80% power calculated for binary combinations of treatment arms on binarized discharge mRS scores

## Discussion

Our findings demonstrate the strength of simulation-based design and power calculation for randomized trials of anti-seizure treatment of EA. This work overcomes limitations of prior trials in the field by defining a data-driven null hypothesis and demonstrating the feasibility and practicality of varying trial designs. We propose our approach as a general recipe for sample size calculations for future trials of anti-seizure treatment of EA in patients with acute brain injuries, to ensure that only high-powered and practical trials that have acceptable likelihood of producing clinically meaningful and applicable results are conducted.

The sample sizes determined by our simulations are significantly larger than those of prior trials examining EEG-guided antiseizure treatments.^16,17^ A recent trial comparing suppression of EA for at least 48 hours vs. standard of care in patients with cardiac arrest and anoxic brain injury found no difference in neurologic outcomes.^17^ 172 subjects (84 per group) were included in the study. The sample size needed was determined based on an assumed prevalence of poor outcomes of 99% extrapolated from other external observational cohorts where poor outcome ranged from 90 to 100 %.^17,29,30^ The investigators specifically aimed at a 7% lower incidence of poor outcomes in the treatment group compared with the control group. This approach to power calculations did not protect them against possible misspecifications of the null (e.g. if 90% were used as the standard of care mortality and a 7% reduction were studied, then the standard difference in proportions power calculations would suggest sample sizes that are more than twice their original recommendations). Eventually their observed data did not match the experimental null hypothesis: The observed proportion of poor outcomes was 90% in their treatment group and 92% in their control group. A second trial comparing lacosamide to phenytoin for the treatment of non-convulsive seizures enrolled only 37 subjects per arm for a non-inferiority trial.^16^ In the absence of efficacy data for anti-seizure medications, the sample size determination was based on consensus from investigators, by contrast to our data-driven methodology. A sample of 200 subjects was deemed as a feasible enrollment target, with power calculations performed for different potential response rates in both arms. The trial enrolled 74 subjects (37 per arm) before the withdrawal of funding.^16^ Generally, it appears that these studies are underpowered to detect the effects they are interested in. Both studies come to a null conclusion that there is no evidence of a difference between the study arms. However, in light of the limitations in defining the null as detailed above, cautious interpretation is indicated for any clinical decision making^31,32^

Using our PK/PD models, we included drug effectiveness in our simulations. Not surprisingly, trials comparing the drug to placebo required the smallest sample sizes, although these sample sizes were still significantly larger than prior trials. There is compelling evidence that EA causes worse outcomes^2^, and therefore trials using a placebo are likely to be considered unethical. There remains significant uncertainty on the optimal treatment approach with multiple treatment strategies employed in current clinical practice ranging from monotherapy with low-dose ASM to combination therapy with high-dose ASMs and anesthetics.^13,33^ In our simulated trials, we considered a few of these treatment strategies and demonstrate that drug choice, dose, and effectiveness all strongly impact trial design. Further PK/PD modeling of other treatment approaches used in clinical practice is indicated to define the full range of treatment responses and a reasonable null model.

A potential explanation for the small effect of anti-epileptiform treatment under our model is that these treatments alone may not be the most effective intervention for aSAH patients with EA. Recall that all patients in the simulation receive standard of care in terms of other treatments.

Development of epileptiform activity in aSAH patients, particularly high-burden epileptiform activity, is also a harbinger of delayed cerebral ischemia (DCI).^34^ In addition, epileptiform activity is associated with increased cerebral metabolism, decreased brain tissue oxygenation, and secondary brain injury.^35^ Therefore, a combination of ASM and/or anesthetic treatment with interventions geared towards increasing cerebral blood flow may need to be investigated in randomized trials of EEG-guided treatment in aSAH. Because functional outcomes are the result of multiple causes, it may also be appropriate and clinically relevant to investigate more proximal outcome measures comprised of EEG findings, clinical exams, and biomarkers of cerebral metabolism in this patient population.

## Limitations

There are several limitations to our study. First, the model for EEG simulations is based on data from a single center, potentially limiting generalizability. We developed one-compartment PK/PD models from a relatively small cohort of patients, and studies are warranted to determine PK/PD and drug effectiveness for different ASMs and anesthetics in the aSAH population with varying disease severity. Our simulations include interventions geared towards increasing cerebral blood flow or metabolism only through standard of care, thus the effects of these interventions are not being directly investigated in our simulated randomized trials. We evaluated discharge functional outcomes, whereas greater effect sizes might be seen with more long-term outcomes (e.g., at 3, 6, or 12 months). Time to randomization in our simulations was restricted to a median of 24 hours (the observed time to EEG initiation in our cohort), and we did not examine the impact of delays or longer time windows to randomization. Early intervention is relevant from a clinical standpoint given evidence that a higher epileptiform burden is associated with worse outcomes in acute brain injury patients.^1^ Finally, we combined all measured EAs into a single EA burden in our trial and did not distinguish between EA subtypes.

In conclusion, our simulated experiments using clinical observational data demonstrate that randomized trials of EEG-guided anti-seizure treatment are currently infeasible. Further data is needed to define the natural course of epileptiform activity in a larger cohort of acute brain injury patients, describe the electrographic response of epileptiform activity to ASMs, and the appropriate therapeutic targets (e.g., EEG alone vs. EEG and biomarkers of metabolism). Further work is also indicated to determine which patient clinical profiles (disease type, severity, metabolism, and EA characteristics) are most likely to have an electrographic and clinical response to ASMs, and whether the response increases with a combination of therapeutic interventions (e.g., ASMs, low dose anesthetics, measures to augment perfusion). Given the clinical complexity of acute brain injury (and in this case aSAH), the dynamic nature of the epileptiform activity, and the multifaceted pathophysiology of secondary brain injury, alternative trial designs with multi-tiered interventions are indicated to determine optimal treatment strategies for EA. Importantly, these future studies fit within the simulation-based RCT paradigm developed in this work that combines modern statistical tools with biologically sound models.

## Acknowledgements

None

## Author contributions

Harsh Parikh contributed to conception and design of the study, analysis of data and drafting a significant portion of the manuscript and figures. Haoqi Sun PhD contributed to conception and design of the study, analysis of data and drafting the manuscript. Rajesh Amerineni contributed to acquisition and analysis of data and drafting the manuscript. Eric S. Rosenthal contributed to acquisition of data and drafting the manuscript. Alexander Volfovsky contributed to conception and design of the study, analysis of data and drafting the manuscript. Cynthia Rudin contributed to conception and design of the study, analysis of data and drafting the manuscript. M. Brandon Westover contributed to conception and design of the study, analysis of data drafting the manuscript. Sahar F. Zafar contributed to conception and design of the study, acquisition and analysis of data and drafting a significant portion of the manuscript and figures.

## Data Availability

The data and software to reproduce these findings are available in a publicly accessible repository at bdsp.io..

